# Chronic Kidney Disease Mineral and Bone Disorder (CKD-MBD) Medication Titration Practices in Hemodialysis Patients in the US

**DOI:** 10.64898/2026.08.02.26359426

**Authors:** Julia J. Scialla, Alyssa Platt, Jonathan Wilson, Rasheeda Hall, Patti L. Ephraim, Daniel E Weiner, L. Ebony Boulware, Jane Pendergast

**Affiliations:** Departments of Medicine and Public Health Sciences, University of Virginia School of Medicine, Charlottesville, VA; Department of Biostatistics and Bioinformatics, Duke University School of Medicine, Durham, NC; Department of Medicine, Duke University School of Medicine, Durham, NC; Advocate Aurora Research Institute, Charlotte, NC; Department of Medicine, Tufts Medical Center, Boston, MA; Wake Forest University School of Medicine, Winston-Salem, NC

**Keywords:** Calcimimetic, cinacalcet, vitamin D, phosphorus, pharmacoepidemiology

## Abstract

Vitamin D sterols, phosphorus binders and calcimimetics are used to treat chronic kidney disease mineral and bone disorder (CKD-MBD) in hemodialysis. With few randomized trials, providers may titrate agents differently reflecting equipoise and opportunities for clinical trials. We studied patients initiating in-center hemodialysis at Dialysis Clinic, Inc facilities from 2006-2015 and who remained on hemodialysis for ≥90 days (n=23,549). Multinomial logit models assessed titration among users of each medication at the start of the month considering static and dynamic CKD-MBD laboratories. Similarly parameterized logistic models assessed treatment initiation. Differences across facilities were quantified as random effects and corresponding median odds ratios. We observed patterns of titration associated with CKD-MBD laboratories including albumin-corrected serum calcium (Ca), serum phosphorus and parathyroid hormone (PTH) and minimal impact of patient characteristics. Best fit models incorporated 3 months of lagged Ca and phosphorus values and linear splines for current Ca, phosphorus and PTH values. Absolute titration probabilities for vitamin D sterols and calcimimetics were influenced by all three CKD-MBD parameters, such that Ca and phosphorus values altered the threshold PTH at which escalation and de-escalation probabilities crossed. Median odds ratios indicated the greatest facility variation for vitamin D sterol titration. Providers titrate CKD-MBD medications based largely on the full CKD-MBD laboratory phenotype, including the recent serum Ca, phosphorus and PTH history. Facility variation suggests equipoise in titration of vitamin D sterols with opportunities for clinical trials.

## Introduction

Medications are widely used to control chronic kidney disease mineral and bone disorder (CKD-MBD) in patients with kidney failure on hemodialysis.^1^ Providers have a choice of agents including: phosphorus binders to limit gastrointestinal phosphorus absorption; vitamin D sterols to replace low levels of active vitamin D and provide feedback inhibition of parathyroid hormone (PTH); and calcimimetics to directly inhibit the release of PTH. Often these medications are used in combination.^2^ Clinical trials of CKD-MBD medications have been primarily industry-sponsored and oriented toward drug approval based on achievement of biochemical control of CKD-MBD laboratories, such as calcium, phosphorus and PTH.^1^ Because better control is *associated* with better overall survival, cardiovascular, and bone outcomes, these drugs are widely used in clinical practice.^3–7^ Observational studies support that use of CKD-MBD medications associates with better clinical outcomes as well.^8^ ^9,10^

Despite compelling observational data, few placebo-controlled or comparative effectiveness clinical trials have been powered for clinical events as the primary outcome. Among well-powered studies, cross-over and dropout has been high, with subsequent results inconclusive.^11,12^ Clinical trials powered for important outcomes and with a high rate of protocol adherence are critically needed to determine the optimal management strategies for CKD-MBD in patients on hemodialysis. Designing clinical trial protocols that mimic widely-held dynamic treatment practices while leveraging areas of practice variability may make trial adherence more achievable. To design these protocols a deeper understanding of practical, but varied, CKD-MBD treatment approaches in real world settings is needed.

In this study, we evaluate titration of CKD-MBD medications within a large population of patients with kidney failure treated with in-center hemodialysis. We aimed to identify clinical factors that associate with subsequent titration of vitamin D sterols, calcimimetics, and phosphorus binders, as well as areas of greater practice variation and equipoise that could be subject to randomization in a trial.

## Methods

### Study Population, Censoring, and Longitudinal Data Organization

This study was conducted using electronic health record (EHR) data for patients initiating in-center hemodialysis at facilities affiliated with Dialysis Clinic, Inc (DCI) between 2006 and 2015. DCI is a medium sized not-for-profit dialysis organization in the US. Patient level DCI EHR data were linked to the United States Renal Data System (USRDS) Standard Analysis Files (SAFs) for additional information on comorbidity, insurance status and clinical outcomes that resulted in censoring, such as dialysis modality changes, death, and kidney transplantation.^13^ The study was approved by DCI, the USRDS, and the Duke University Institutional Review Board.

Included participants remained on in-center hemodialysis at a DCI facility 90 days after their incident hemodialysis date. Patients were permanently censored at the time of death, kidney transplantation, permanent transfer out of DCI facilities, or on December 31, 2015. Observations were organized into person-months based on exact calendar months, beginning at the first complete month 90-days after starting hemodialysis. A patient-month was censored if model covariates were missing, there were no in-center hemodialysis treatments at the DCI facility during the month, or if a modality other than in-center hemodialysis was used during the month. Among the CKD-MBD medications available during the time of study, vitamin D sterols could have intravenous or oral routes of administration. In order to allow the dose to stabilize with the new route, participants were censored from vitamin D sterol titration models during months in which a route change was observed.

### Medication Ascertainment and Classification

Use of vitamin D sterols, calcimimetics, and phosphorus binders was ascertained from the DCI Medication List using Generic Product Identifier (GPI) codes, as previously described.^2^ Doses of vitamin D sterols were converted to paricalcitol equivalents by multiplying doses by 4 for calcitriol and 1.6 for doxercalciferol.^14–16^ During the study period, cinacalcet was the only available calcimimetic. Drug doses were converted to daily doses based on their frequency. We classified phosphorus binders as either low range or high range and then generated an ordinal variable for modeling dose titration: no binder; 1 low dose binder; 1 high dose binder; 2 or more binders at any doses (**Supplemental Methods**). ‘Ceiling doses,’ at which the drug may not be able to be titrated further, were considered *a priori* to be 120 mg daily for cinacalcet, 36 mcg weekly for paricalcitol equivalents, and 2 or more phosphorus binders at any dose. These were quantified to determine impact on model interpretation.

### Outcomes

Our primary outcome was a multinomial classification, indicating (i) a decrease or discontinuation (de-escalation), (ii) an increase in medication dose (escalation), or (iii) no change in dosage, the reference category. If more than one change occurred in a month, we chose the first change. Because patients who were not using a medication were not eligible for de-escalation, inclusion in the model was conditional on having an active medication at the start of the month. Secondary models evaluated drug initiation among non-users at the start of the month.

### Predictors of Medication Titration and Other Covariates

Covariates of interest included demographics, comorbid health conditions and health status, medication insurance coverage, time since dialysis initiation, calendar year, dialysis adequacy and duration, dialysate calcium concentration, and recent CKD-MBD laboratories including serum phosphorus, albumin-corrected serum calcium and PTH (**Supplemental Methods**). Covariates tested for influence on titration models included age categories (<30, 30-39, 40-49, 50-59, 60-64, 65-69, 70-74, 75-80, 80-84, ≥85 years), sex, diabetes, total comorbidity score (0-1, 2-3, 4-6, 7-9, ≥10), functional limitation, and CKD-MBD laboratories.

According to standard dialysis practice, serum phosphorus and albumin-corrected serum calcium were typically measured monthly and PTH quarterly. However, in any given person-month more than 1 set of laboratories may be drawn. In each case we selected the most recent laboratories up to and including the date of the index medication change or the first set of laboratories in the month if there were no medication changes. Because many providers also evaluate trends in laboratories, we included lagged parameters for serum phosphorus and calcium quantified as difference between current and prior values over the last 2 months. A detailed schematic is depicted in the **Supplemental Methods.**

### Statistical Analysis

We used a mixed effects multinomial logistic regression model to estimate the conditional probability of de-escalation, escalation, or no change of each medication (**Supplemental Methods**). Based on model fit, we included linear spline functions with cutpoints at PTH 300 and 400 pg/ml, serum phosphorus at 5.5 mg/dl, and serum calcium of 10.2 mg/dl were incorporated in all models. An additional cutpoint for serum phosphorus at 7 mg/dl was incorporated for models of phosphorus binders. Addition of patient characteristics including age categories (<30, 30-39, 40-49, 50-59, 60-64, 65-69, 70-74, 75-80, 80-84, ≥85 years), sex, diabetes, total comorbidity score (0-1, 2-3, 4-6, 7-9, ≥10), and functional limitation were also evaluated for fit (**Supplemental Methods**). For parsimony, patient characteristics were not included in final models.

To model practice differences across facilities and individuals, we included random intercepts in the models for each dialysis facility and each individual. Including random intercepts for individuals was found to be problematic and unnecessary, due to near-zero variability in the outcome probabilities across individuals when laboratory structures were included in the models. Because differences in laboratories accounted for most of the differences in management across patients and periods, periods were treated as functionally independent observations.

Clinic-to-clinic variability was expressed as median odds ratios, rather than intraclass correlation coefficients (ICCs), for easier interpretation. Median odds ratios reflect the median of the distribution of odds ratios generated for each randomly chosen pair of facilities with the higher odds always in the numerator. These can be interpreted as the relative difference in predicted odds for identical individuals treated in different facilities, and thus is a measure of practice variation. Confidence intervals were produced using Wald confidence limits around the variance of the random intercepts.

We qualitatively assessed model fit by comparing model-based mean conditional predicted probabilities for the sample to the model-free 95% exact confidence intervals around observed corresponding probabilities. Model results were displayed according to panels depicting all 3 CKD-MBD laboratories in concert, consistent with prior studies depicting the clinical relevance of laboratory combinations, or ‘phenotypes’.^3,17–19^

Secondary models evaluated initiation of each drug class with the same parameterization. All analyses were conducted using SAS 9.4 with model assessments using the GLIMMIX procedure.

## Results

### Study Population Characteristics and Cohort Flow

We identified 31,111 patients who initiated in-center hemodialysis for kidney failure at a DCI facility between 2006 and 2015. A total of 23,549 participants remained in the sample and were eligible at 90 days (**Supplemental Figure 1**). Patients could contribute person-months to multiple models based on having an eligible person-month.

Ultimately, in titration models, 12,889 unique individuals had at least 1 person-month included for vitamin D sterols, 5,179 for calcimimetics, and 17,071 for phosphorus binders. Patients included in the calcimimetic titration models were younger, more likely to be Black, had higher PTH levels, had lower comorbidity, and were of later dialysis vintage when entering the sample compared to the other titration study populations (**Table 1**). More of these patients left the sample due to censoring at the end of the observation period compared with populations for other titration models where death was the major reason for censoring. Characteristics of participants in secondary models of medication initiation are depicted in **Supplemental Table 1**.

**Table 1.** Baseline Characteristics of Titration Model Study Populations.

| Median, Interquartile range or n(%) | Calcimimetic<br>(N=5,179) | Phosphorus Binder<br>(N=17,071) | Vitamin D Sterol<br>(N=12,889) |
| --- | --- | --- | --- |
| **Age at incidence (years)** | 59<br>48, 68 | 63<br>52, 72 | 62<br>52, 72 |
| **Race** |  |  |  |
| White | 2,322 (44.8%) | 9,977 (58.4%) | 6,887 (53.4%) |
| Black/African American | 2,671 (51.6%) | 6,273 (36.7%) | 5,441 (42.2%) |
| Other | 186 (3.6%) | 821 (4.8%) | 561 (4.4%) |
| **Sex** |  |  |  |
| Male | 2,728 (52.7%) | 9,669 (56.6%) | 7,157 (55.5%) |
| Female | 2,451 (47.3%) | 7,402 (43.4%) | 5,732 (44.5%) |
| **Hispanic ethnicity** |  |  |  |
| No | 4,853 (93.7%) | 15,976 (93.6%) | 12,005 (93.2%) |
| Yes | 324 (6.3%) | 1,086 (6.4%) | 875 (6.8%) |
| **Primary cause of kidney failure** |  |  |  |
| Diabetes | 2,349 (48.6%) | 8,087 (51.1%) | 6,070 (51.0%) |
| Hypertension | 1,536 (31.8%) | 4,596 (29.1%) | 3,627 (30.5%) |
| Glomerulonephritis | 460 (9.5%) | 1,366 (8.6%) | 957 (8.0%) |
| Other cause | 486 (10.1%) | 1,764 (11.2%) | 1,239 (10.4%) |
| **Comorbidity index** | 2<br>1, 4 | 3<br>1, 5 | 3<br>1, 5 |
| **Previously diagnosed with diabetes (CMS Form 2728)** |  |  |  |
| No | 2,280 (44.1%) | 6,892 (40.4%) | 5,206 (40.4%) |
| Yes | 2,893 (55.9%) | 10,157 (59.6%) | 7,665 (59.6%) |
| **Inability to ambulate** |  |  |  |
| No | 4,713 (95.4%) | 15,282 (93.8%) | 11,579 (94.5%) |
| Yes | 229 (4.6%) | 1,007 (6.2%) | 669 (5.5%) |
| **Parathyroid hormone (90 days; pg/mL)** | 336<br>182, 537 | 252<br>138, 441 | 268<br>152, 448 |
| **Phosphorus (90 days; mg/dL)** | 5.5<br>4.6, 6.7 | 5.3<br>4.4, 6.4 | 5.2<br>4.4, 6.3 |
| **Albumin corrected calcium (90 days; mg/dL)** | 9.4<br>9.0, 9.8 | 9.3<br>8.9, 9.7 | 9.3<br>8.9, 9.7 |
| **Serum albumin (90 days; g/dL)** | 3.7<br>3.4, 4.0 | 3.7<br>3.4, 3.9 | 3.7<br>3.4, 3.9 |
| **Dialysis time (90 days; min/week)** | 675<br>630, 720 | 630<br>630, 720 | 630<br>630, 720 |
| **Kt/V (90 days)** | 1.5<br>1.4, 1.7 | 1.5<br>1.4, 1.7 | 1.5<br>1.4, 1.7 |
| **Dialysate calcium (90 days; mEq/L)** | 2.5<br>2.5, 2.5 | 2.5<br>2.5, 2.5 | 2.5<br>2.5, 2.5 |

From these unique individuals, 263,093 person-months were included for titration of vitamin D sterols, 95,356 for titration of calcimimetics, and 426,891 for titration of phosphorus binders. Vitamin D sterols were titrated most frequently with 45,653 months characterized by de-escalation (17.4%) and 33,246 months characterized by escalation (12.6%). Calcimimetics and phosphorus binders were less frequently titrated (<5% of months for each; **Supplemental Table 2**). Calcimimetics were also less frequently initiated (1.6% of months) compared with vitamin D sterols (10.1%) and phosphorus binders (8.0%).

### CKD-MBD Laboratories by Titration Status of Medications

We first evaluated the distribution of key CKD-MBD laboratories in the current and lagged months across outcomes (**Figure 1**). PTH was higher on average in patient-months in which vitamin D or calcimimetics were escalated and lower on average in months in which vitamin D or calcimimetics were de-escalated, with each compared with no change (p<0.01 for each; **Figure 1a**). Serum calcium was higher on average among those who de-escalated vitamin D sterols compared to those with no change (p<0.01; **Figure 1b**). Serum phosphorus was higher on average in patient-months in which vitamin D sterols were de-escalated, calcimimetics were escalated, or phosphorus binders were escalated, each compared to no change (each p<0.01; **Figure 1c**). Examining the lagged laboratories revealed a consistent picture with evidence for directionally similar patterns with the preceding months’ dynamic laboratories (i.e., rising or falling; **Table 2**). We observed overall similar patterns for the initiation outcomes (**Supplemental Table 3**).

**Figure 1.**
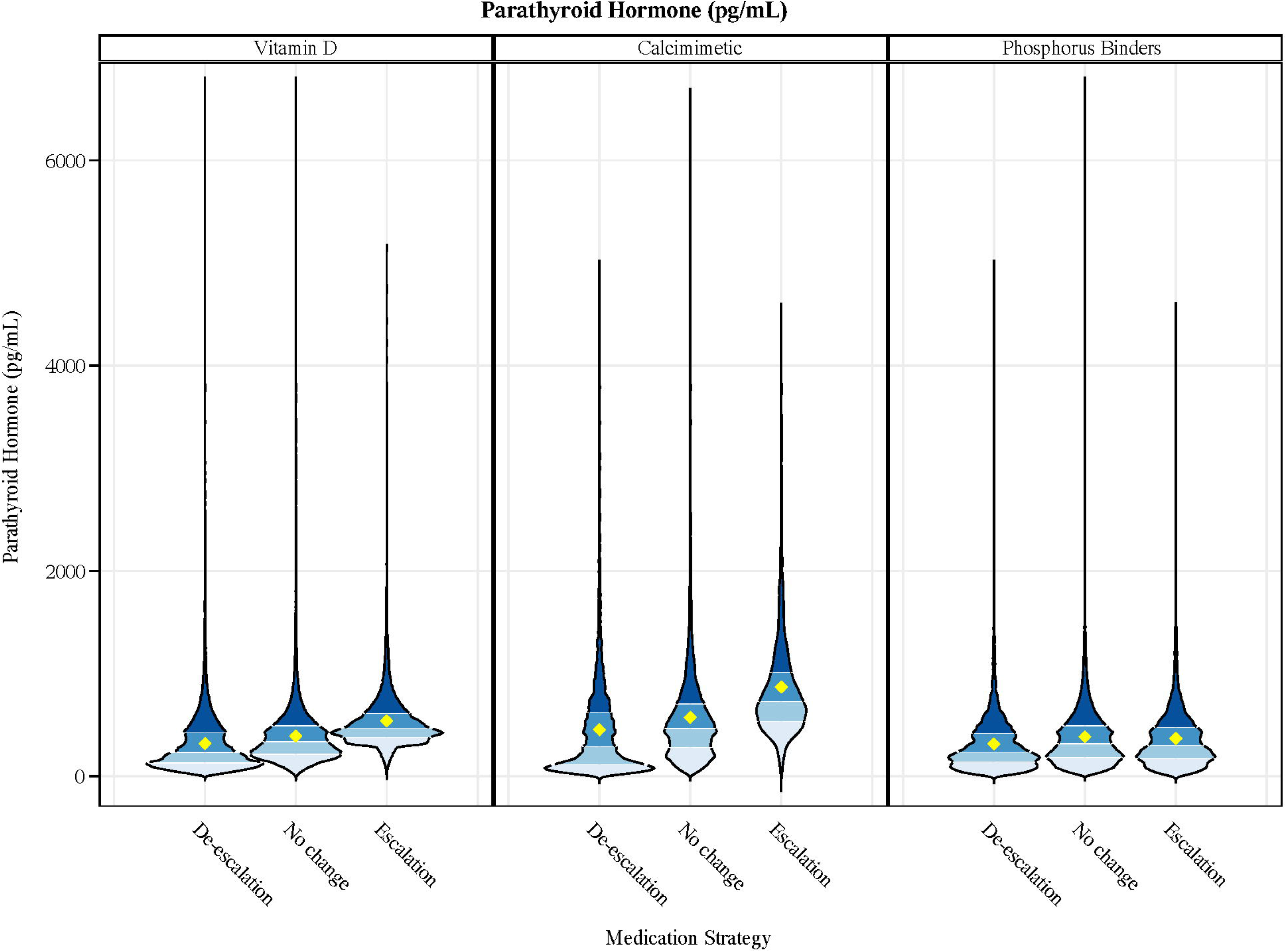
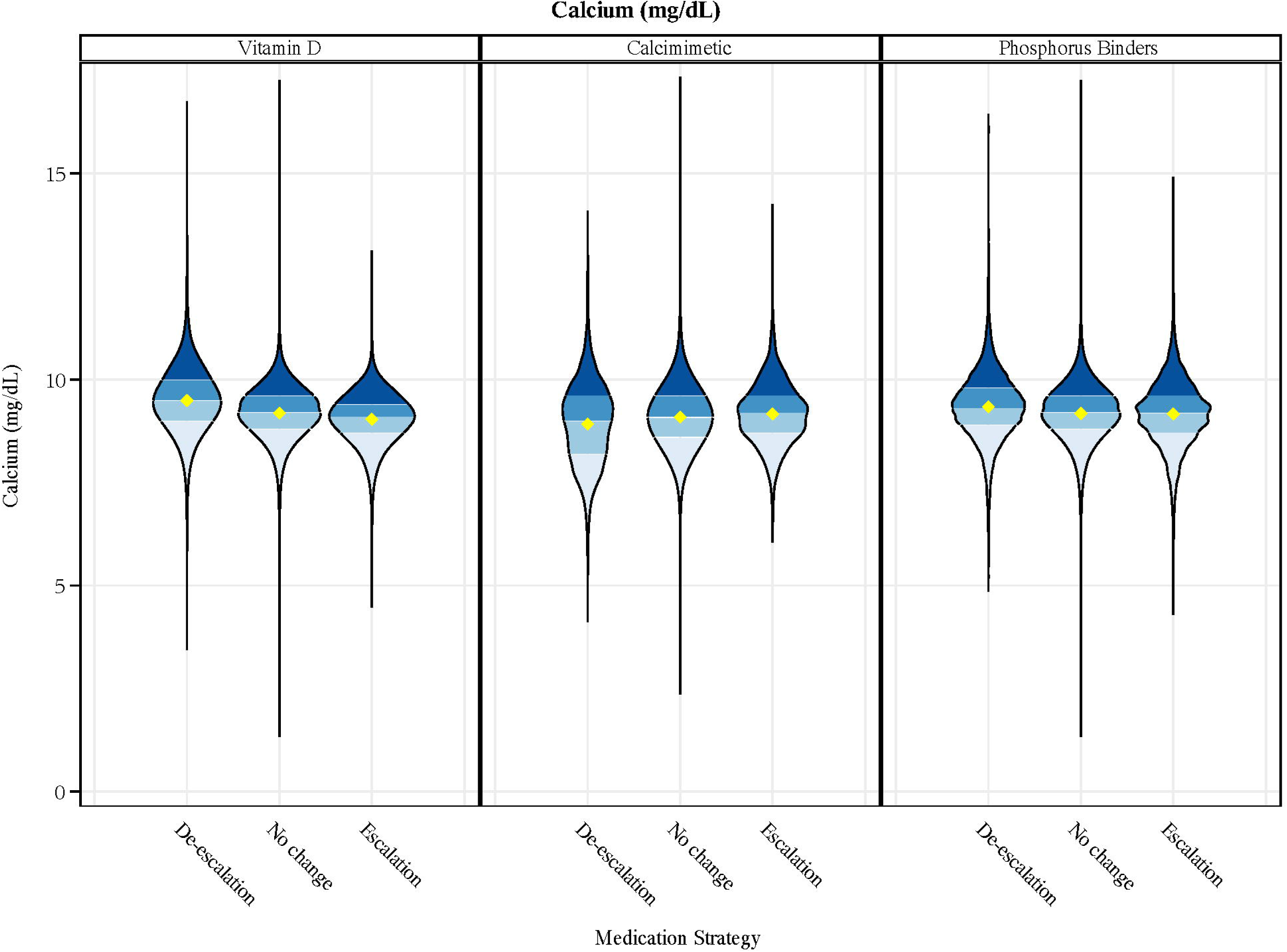
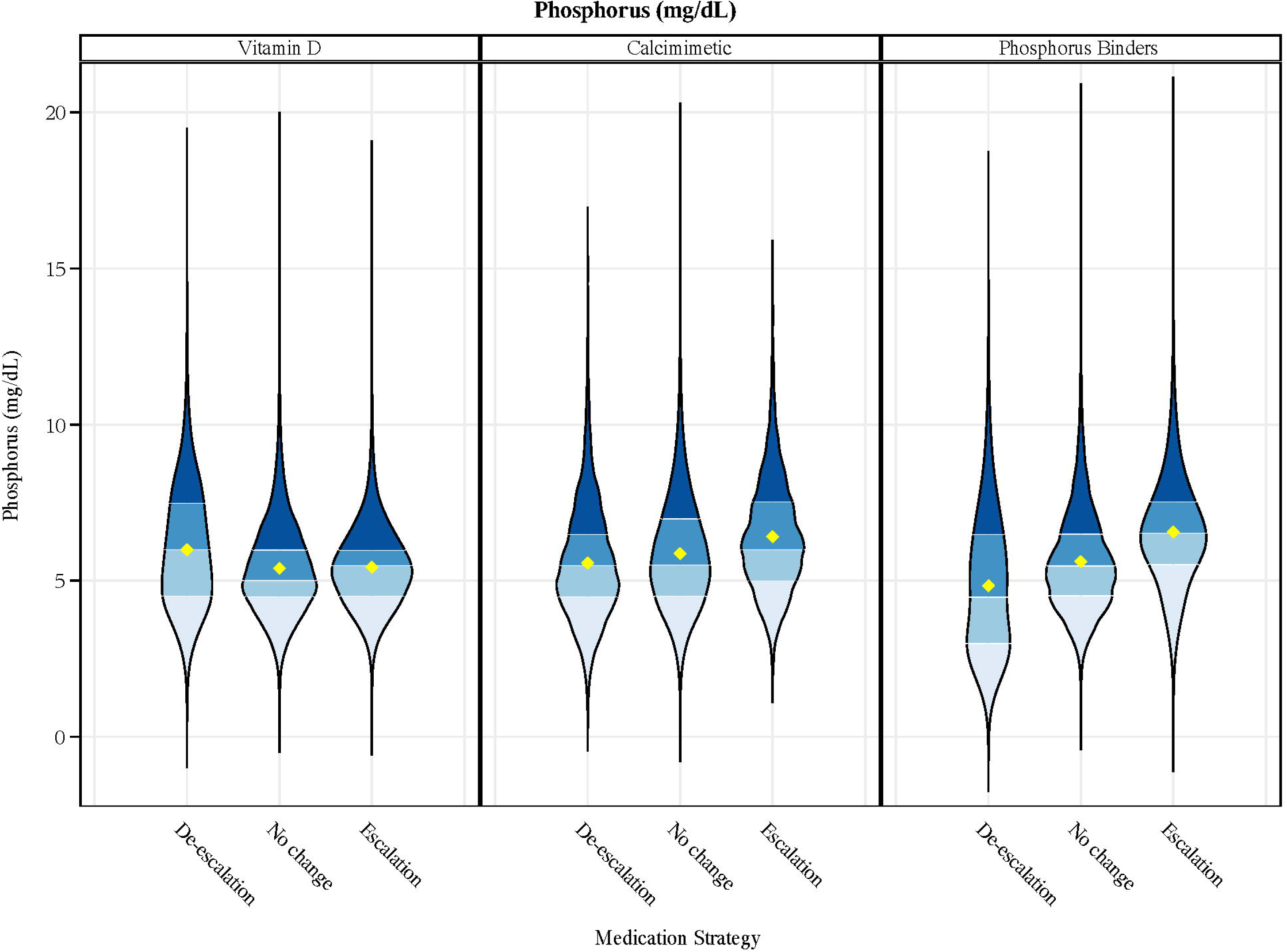
Values of CKD-MBD Laboratories by Categories of CKD-MBD Medication Titrations. Panel A depicts serum parathyroid hormone levels in pg/ml. Panel B depicts albumin-corrected serum calcium levels in mg/dl. Panel C depicts serum phosphorus in mg/dl. The distribution of each laboratory is presented across categories of de-escalation, no change and escalation for each medication (vitamin D sterols, calcimimetics and phosphorus binders) using violin plots. The width of the ‘violin’ at a particular point is proportional to the density of measurements at that value. Diamonds depict the mean and differences in shading depict the quartiles. ‘Violins’ extend the full range of the data from the lowest to highest values, including any outliers. Serum calcium was rounded to the nearest 0.1 mg/dl and serum phosphorus to the nearest 0.5 mg/dl for these plots for smoothing.

**Table 2.**
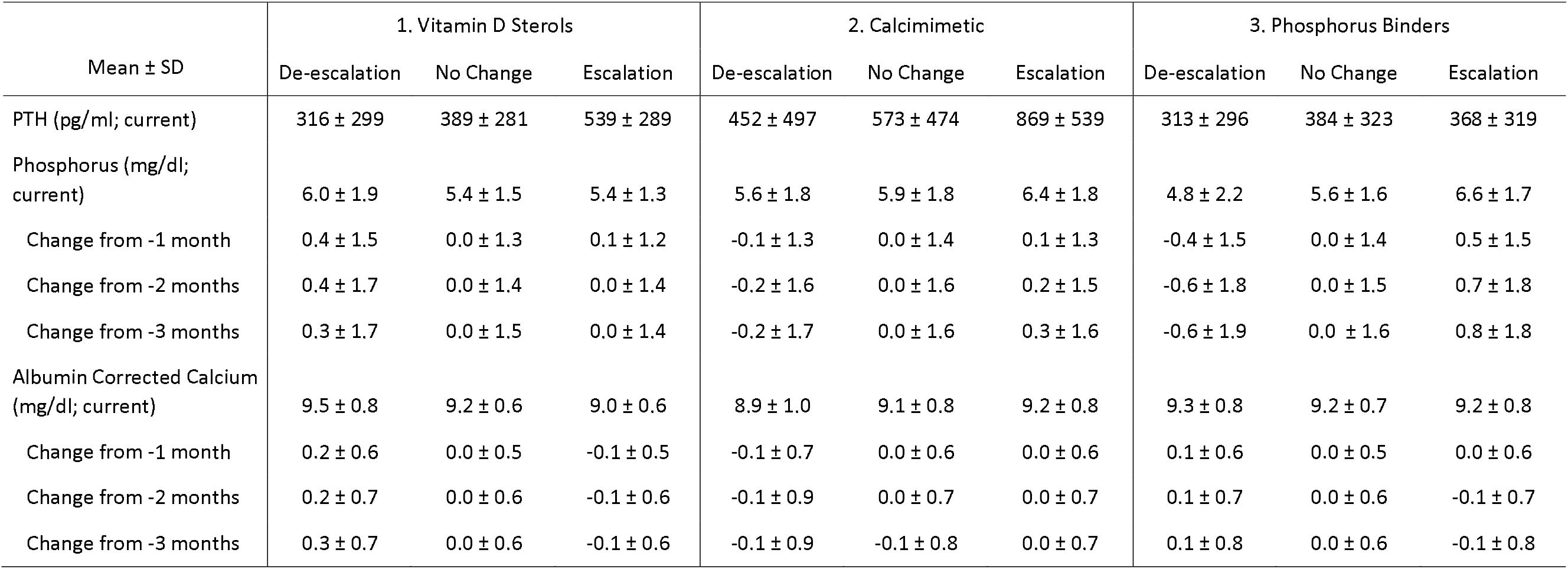
Current and Lagged Laboratory Values by Titration Status of for Vitamin D Sterols, Calcimimetics, and Phosphorus Binders.

### Models of CKD-MBD Medication Titration Based on Laboratory Phenotypes

We next used multivariable models and graphic displays stratified by panels of laboratories to depict the relationship between integrated laboratory phenotypes and subsequent drug titration. Addition of lagged laboratories to spline models meaningfully improved model fit by Bayesian Information Criterion. Compared to the impact of laboratories, addition of baseline patient characteristics did not, or only marginally, improved model fit compared to the impact of dynamic laboratories (**Supplemental Table 4**).

Depictions of the absolute modeled probability of each CKD-MBD medication titration according to the full set of CKD-MBD laboratories is displayed in **Figures 2-4** to facilitate clinical interpretation. Vitamin D sterols (**Figure 2**) were more likely to be de-escalated when serum calcium is high (**Figure 2, top row**), and more likely to be escalated as PTH rises above approximately 300 pg/ml when serum calcium is normal (**Figure 2, middle row**). The association of higher PTH and vitamin D sterol escalation was less pronounced when serum phosphorus was high, but the PTH value at which the probability of escalating exceeds the probability of de-escalating (i.e. point at which red and blue lines cross) is lower when serum calcium is lower (**Figure 2, bottom row**).

**Figure 2.**
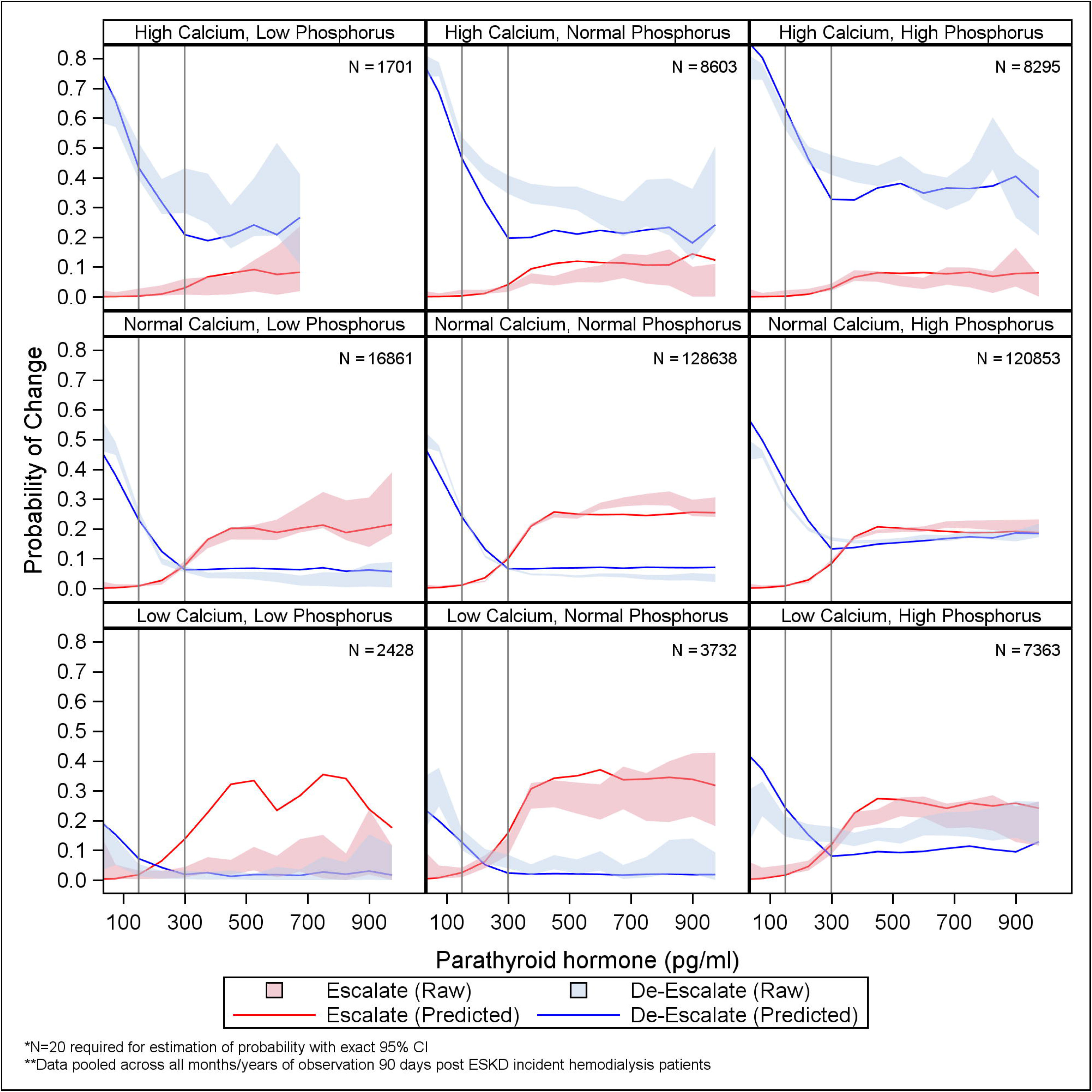
Model Predicted vs. Observed Raw Probability of Titration of Vitamin D Sterols Based on Laboratory Panels. Results are derived from a mixed effects multinomial logistic regression model to estimate the conditional probability of de-escalation, escalation, or no change of vitamin D sterols among users at the start of the month as a function of CKD-MBD laboratories. Models include linear spline functions for each laboratory with cutpoints at PTH 300 and 400 pg/ml, serum phosphorus at 5.5 pg/ml, and serum calcium at 10.2 mg/dl, 3 sets of lagged laboratories for serum calcium and phosphorus, and random effects for facilities. Models pool across all person-months of data. Red lines demonstrate the model predicted probability of escalation across increasing values of serum PTH for those with serum calcium and serum PTH in the described range, where low calcium is <8.0 mg/dl, normal calcium is 8.0-10.2 mg/dl and high calcium is >10.2 mg/dl, and low phosphorus is < 3.5 pg/dl, normal phosphorus is 3.5-5.5 mg/dl and high phosphorus is >5.5 mg/dl. Blue lines demonstrate the model predicted probability of de-escalation across increasing values of serum PTH in similar panels. Probability of ‘no-change’ is not depicted but is implied because categories are mutually exclusive and exhaustive. Lines are truncated when less than 20 person/months contribute to the calculation of means. Bands depict 95% exact binomial confidence intervals around each line.

Calcimimetics were generally more likely to be escalated compared to de-escalated when PTH was greater than 300 pg/ml except when serum calcium or phosphorus were low **(Figure 3**). Phosphorus binders were rarely de-escalated when serum phosphorus was above 5.5 mg/dl, with the possible exception of when PTH was low and serum calcium was high. Probability of escalating phosphorus binders began to rise around 5.5 mg/dl (**Figure 4**). Lagged laboratories were associated with medication titrations independent of the current laboratories in a manner that was directionally similar to unmodeled results (**Supplemental Table 5**).

**Figure 3.**
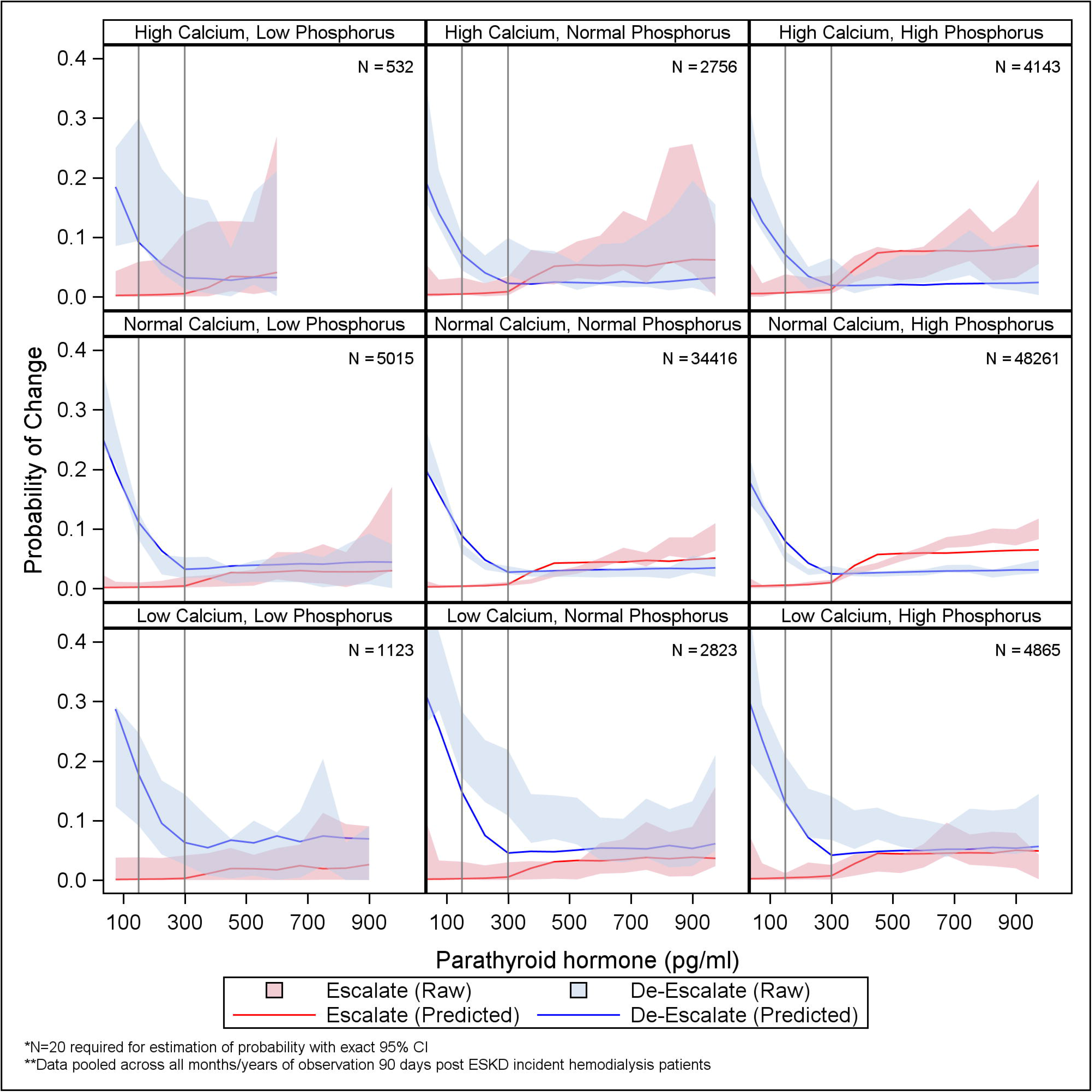
Model Predicted vs. Observed Raw Probability of Titration of Calcimimetics Based on Laboratory Panels. Results are derived from a mixed effects multinomial logistic regression model to estimate the conditional probability of de-escalation, escalation, or no change of calcimimetics among users at the start of the month as a function of CKD-MBD laboratories. Models include linear spline functions for each laboratory with cutpoints at PTH 300 and 400 pg/ml, serum phosphorus at 5.5 pg/ml, and serum calcium at 10.2 mg/dl, 3 sets of lagged laboratories for serum calcium and phosphorus, and random effects for facilities. Models pooling across all person-months of data. Red lines demonstrate the model predicted probability of escalation across increasing values of serum PTH for those with serum calcium and serum PTH in the described range, where low calcium is <8.0 mg/dl, normal calcium is 8.0-10.2 mg/dl and high calcium is >10.2 mg/dl, and low phosphorus is < 3.5 pg/dl, normal phosphorus is 3.5-5.5 mg/dl and high phosphorus is >5.5 mg/dl. Blue lines demonstrate the model predicted probability of de-escalation across increasing values of serum PTH in similar panels. Probability of ‘no-change’ is not depicted but is implied because categories are mutually exclusive and exhaustive. Lines are truncated when less than 20 person/months contribute to the calculation of means. Bands depict 95% exact binomial confidence intervals around each line.

**Figure 4.**
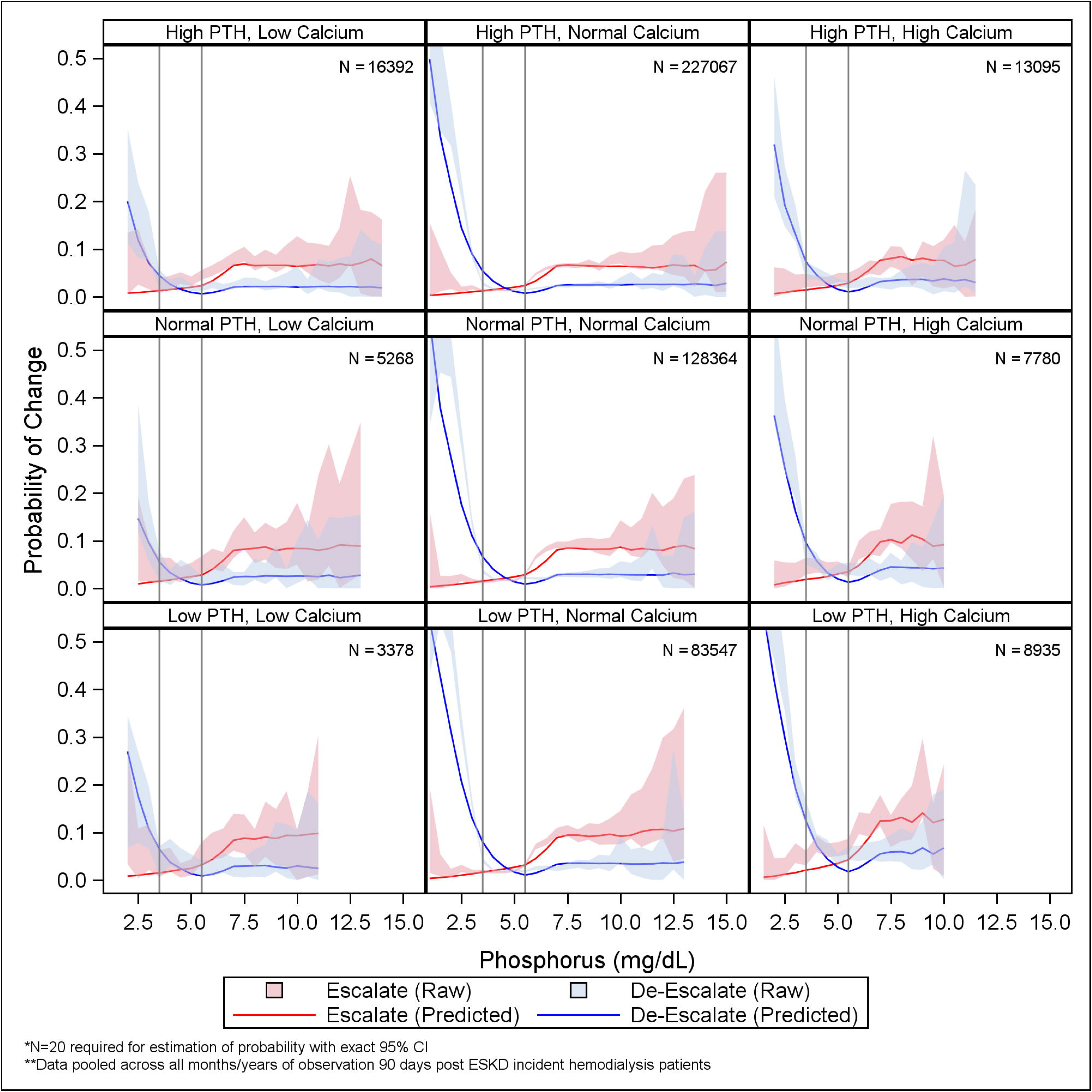
Model Predicted vs. Observed Raw Probability of Titration of Phosphorus Binders Based on Laboratory Panels. Results are derived from a mixed effects multinomial logistic regression model to estimate the conditional probability of de-escalation, escalation, or no change of phosphorus binders among users at the start of the month as a function of CKD-MBD laboratories. Models include linear spline functions for each laboratory with cutpoints at PTH 300 and 400 pg/ml, serum phosphorus at 5.5 and 7.0 mg/dl, and serum calcium at 10.2 mg/dl, 3 sets of lagged laboratories for serum calcium and phosphorus, and random effects for facilities. Models pool across all person-months of data. Red lines demonstrate the model predicted probability of escalation across increasing values of serum phosphorus for those with serum calcium and serum PTH in the described range, where low PTH is <150 pg/ml, normal PTH is 150-300 pg/ml and high PTH is >300 pg/ml and low calcium is <8.0 mg/dl, normal calcium is 8.0-10.2 mg/dl and high calcium is >10.2 mg/dl. Blue lines demonstrate the model predicted probability of de-escalation across increasing values of serum phosphorus in similar panels. Probability of ‘no-change’ is not depicted but is implied because categories are mutually exclusive and exhaustive. Lines are truncated when less than 20 person/months contribute to the calculation of means. Bands depict 95% exact binomial confidence intervals around each line.

CKD-MBD laboratory effects on medication initiation were similar (**Supplemental Table 6**). Few patient months were characterized by medications at our pre-specified ceiling doses for vitamin D sterols or calcimimetics (**Supplemental Table 7**). Given the ordinal nature of our phosphorus binder dosing categorization, these were frequently at ceiling doses (**Supplemental Table 7**) limiting escalation in some scenarios.

### Practice Variation in CKD-MBD Medication Titration

Facility-level random intercepts suggested heterogeneity in practice that was most marked for vitamin D sterol titration. Compared to the reference of ‘no change’, the median odds ratio for titration of calcimimetics was 1.43 (95% CI 1.37 to 1.53) for escalation and 1.49 (95% CI 1.42 to 1.60) for de-escalation across facilities. For phosphorus binders, the median odds ratio was 1.35 (95% CI 1.30 to 1.40) for escalation and 1.49 (95% CI 1.42 to 1.58) for de-escalation across facilities. For vitamin D sterols the median odds ratio was 1.88 (95% CI 1.77 to 2.02) for escalation and 2.40 (95% CI 2.23 to 2.69) for de-escalation across facilities.

## Discussion

In this study of real-world treatment of CKD-MBD in US hemodialysis patients we note two key findings: 1) prescribers respond to the full constellation of calcium, phosphorus and PTH over the prior 3 months when titrating CKD-MBD medications; and 2) using facilities as proxies of providers, there is substantial heterogeneity in titration, particularly in the approach to titrating vitamin D sterols. Areas of relatively consistent practice (e.g., titration patterns of phosphorus binders and calcimimetics) could guide design of a practical, real-world titration protocol that would be acceptable to most providers and limit cross-over in a future randomized trial. Areas of heterogeneous practice (e.g., titration of vitamin D sterols) are excellent candidates to be randomized in a future comparative effectiveness trial.

Both vitamin D sterols and calcimimetics are indicated for control of secondary hyperparathyroidism in patients with kidney failure on dialysis.^20,21^ We found strong association of higher PTH levels with escalation and lower PTH levels with de-escalation of these medications. However, titration was also strongly influenced by other CKD-MBD laboratories. We found that vitamin D sterols were infrequently titrated at any PTH when serum calcium was high. Higher serum calcium and serum phosphorus were more strongly associated with de-escalation of vitamin D sterols, but escalation of calcimimetics. The opposite patterns were noted when these laboratories were low or falling. These strong patterns in use suggest that providers utilize CKD-MBD medications in an integrated manner to achieve simultaneous biochemical control of not only PTH, but also calcium and phosphorus. This type of integrated management strategy makes physiological sense and maintains guideline concordant laboratories, but has not been shown to improve outcomes. Most providers and guidelines infer improved outcomes from the extensive observational literature linking CKD-MBD laboratory features with survival and cardiovascular disease.

Recently, Danese *et al* demonstrated a stronger association between CKD-MBD laboratories and mortality when the laboratories were evaluated in concert rather than separately.^22^ Our results suggest that providers already interpret laboratories holistically when treating. Smaller trials in CKD-MBD have compared different agents or integrated titration protocols that incorporate multiple agents in the approach to treatment.^23–28^ To date these have been primarily industry sponsored and have not focused on patient-centered or clinical outcomes. Understanding integrated titration approaches used in current practice could help develop larger trials with management strategies that support adherence. Of note, the current models reported here evaluate medication titration of one class at a time. Integrated approaches would need to consider simultaneous titration of different classes and require further development.

Overall, calcimimetics and phosphorus binders were titrated at lower frequency than vitamin D sterols and exhibited lower variation across facilities. At the time of this data, both of these medications were only available orally and largely administered at home. To avoid confusion with patient-administered medications, we propose that providers may be more likely to frequently titrate other medications that are administered at dialysis, like vitamin D sterols. Alternatively, they may first alter other aspects of treatment that we did not model, such as the dialysate calcium. In the case of patient administered medications, providers may focus more heavily on medication reconciliation and supporting adherence than on titration. In addition, poor tolerance of these medications, or ceiling effects, in which maximum doses have been reached in many patients, could limit further escalation. Evaluation of titration of etelcalcetide, in which administration in the clinic is controlled by the provider, could yield different results.

Our study is one of the largest evaluations of provider prescribing responses to CKD-MBD laboratories within in-center hemodialysis. Our data set includes detailed ascertainment of medications and medication doses that were previously validated.^2^ Use of EHR data allows us to evaluate detailed panels of laboratories, which are not available in alternative sources such as the USRDS. We also have limitations. We evaluated provider practices within a single dialysis organization. It is possible that practices could differ in other large dialysis for-profit organizations, independent, or academic organizations. To our knowledge, DCI did not use universal protocols for CKD-MBD therapy at this time, which could allow greater variability than other providers.

In summary, we describe the central importance of the full CKD-MBD laboratory phenotype in CKD-MBD medication titration decisions for patients with kidney failure receiving in-center hemodialysis. We noted important variation across facilities, most notably for the pattern of vitamin D sterol titration. Design of CKD-MBD medication titration algorithms for clinical trials will need to provide guidance based on simultaneous consideration of calcium, phosphorus and PTH with most flexibility in the use of vitamin D sterols.

## Data Availability

The data used in this study were provided by Dialysis Clinic, Inc (DCI) and United States Renal Data System (USRDS). Linked data from DCI and USRDS cannot be directly shared by the study team based on the terms of their data use agreements.

## Acknowledgments

The authors are grateful to the staff and patients of Dialysis Clinic, Inc (DCI). This study was supported by R01DK111952 from the National Institute of Diabetes and Digestive and Kidney Diseases. Additional support was provided in part by National Institute on Aging Award Number K76AG059930 (RH), National Center for Advancing Translational Sciences of the National Institutes of Health Award Number UL1TR002553, and the ASN Foundation for Kidney Research (RH). Neither the sponsors, nor DCI had a deciding role in the study design, analysis, interpretation of the data, writing of the report, or the decision to submit the report for publication. The manuscript reflects the interpretation and opinions of the authors and is not expressly endorsed by the National Institutes of Health, the National Institute of Diabetes and Digestive and Kidney Diseases, National Institute on Aging, or DCI. The data reported here have been supplied by the United States Renal Data System (USRDS). The interpretation and reporting of these data are the responsibility of the author(s) and in no way should be seen as an official policy or interpretation of the U.S. government. Results of these analyses have not been reported previously except in abstract format at the American Society of Nephrology’s 2018 Kidney Week in San Diego, CA.

## References

1. Scialla JJ. Evidence basis for integrated management of mineral metabolism in patients with end-stage renal disease. Curr Opin Nephrol Hypertens. Jul 2018;27(4):258–267. doi:10.1097/MNH.0000000000000417

2. Hall R, Platt A, Wilson J, et al. Trends in Mineral Metabolism Treatment Strategies in Patients Receiving Hemodialysis in the United States. Clin J Am Soc Nephrol. Nov 6 2020;15(11):1603–1613. doi:10.2215/CJN.04350420

3. Tentori F, Blayney MJ, Albert JM, et al. Mortality risk for dialysis patients with different levels of serum calcium, phosphorus, and PTH: the Dialysis Outcomes and Practice Patterns Study (DOPPS). Am J Kidney Dis. Sep 2008;52(3):519–30. doi:10.1053/j.ajkd.2008.03.020

4. Palmer SC, Hayen A, Macaskill P, et al. Serum levels of phosphorus, parathyroid hormone, and calcium and risks of death and cardiovascular disease in individuals with chronic kidney disease: a systematic review and meta-analysis. JAMA. Mar 16 2011;305(11):1119–27. doi:10.1001/jama.2011.308

5. Fernandez-Martin JL, Martinez-Camblor P, Dionisi MP, et al. Improvement of mineral and bone metabolism markers is associated with better survival in haemodialysis patients: the COSMOS study. Nephrol Dial Transplant. Sep 2015;30(9):1542–51. doi:10.1093/ndt/gfv099

6. Fukagawa M, Kido R, Komaba H, et al. Abnormal mineral metabolism and mortality in hemodialysis patients with secondary hyperparathyroidism: evidence from marginal structural models used to adjust for time-dependent confounding. Am J Kidney Dis. Jun 2014;63(6):979–87. doi:10.1053/j.ajkd.2013.08.011

7. Fuller DS, Dluzniewski PJ, Cooper K, Bradbury BD, Robinson BM, Tentori F. Combinations of mineral and bone disorder markers and risk of death and hospitalizations in the international Dialysis Outcomes and Practice Patterns Study. Clinical Kidney Journal. 2020;13(6):1056–1062. doi:10.1093/ckj/sfz112

8. Platt A, Wilson J, Hall R, et al. Comparative Effectiveness of Alternative Treatment Approaches to Secondary Hyperparathyroidism in Patients Receiving Maintenance Hemodialysis: An Observational Trial Emulation. Am J Kidney Dis. Jan 2024;83(1):58–70. doi:10.1053/j.ajkd.2023.05.016

9. Iseri K, Bieber B, Hida N. Calcimimetic and vitamin D receptor agonist therapy associates with lower mortality and fractures in hemodialysis patients: an international DOPPS analysis. Clinical Kidney Journal. 2026;doi:10.1093/ckj/sfag164

10. Isakova T, Gutiérrez OM, Chang Y, et al. Phosphorus binders and survival on hemodialysis. J Am Soc Nephrol. Feb 2009;20(2):388–96. doi:10.1681/asn.2008060609

11. Suki WN, Zabaneh R, Cangiano JL, et al. Effects of sevelamer and calcium-based phosphate binders on mortality in hemodialysis patients. Kidney Int. Nov 2007;72(9):1130–7. doi:10.1038/sj.ki.5002466

12. EVOLVE Trial Investigators, Chertow GM, Block GA, et al. Effect of cinacalcet on cardiovascular disease in patients undergoing dialysis. N Engl J Med. Dec 27 2012;367(26):2482–94. doi:10.1056/NEJMoa1205624

13. United States Renal Data System. 2019 USRDS annual data report: Epidemiology of kidney disease inthe United States. . 2019.

14. Goldenberg MM. Paricalcitol, a new agent for the management of secondary hyperparathyroidism in patients undergoing chronic renal dialysis. Clin Ther. Mar 1999;21(3):432–41. doi:10.1016/S0149-2918(00)88299-5

15. Zisman AL, Ghantous W, Schinleber P, Roberts L, Sprague SM. Inhibition of parathyroid hormone: a dose equivalency study of paricalcitol and doxercalciferol. Am J Nephrol. Nov-Dec 2005;25(6):591–5. doi:10.1159/000089707

16. Kumar J, Tran NT, Schomberg J, Streja E, Kalantar-Zadeh K, Pahl M. Successful Conversion From Parenteral Paricalcitol to Pulse Oral Calcitriol for the Management of Secondary Hyperparathyroidism in Hemodialysis Patients. J Ren Nutr. Jul 2016;26(4):265–9. doi:10.1053/j.jrn.2016.02.006

17. Block GA, Kilpatrick RD, Lowe KA, Wang W, Danese MD. CKD-mineral and bone disorder and risk of death and cardiovascular hospitalization in patients on hemodialysis. Clin J Am Soc Nephrol. Dec 2013;8(12):2132–40. doi:10.2215/CJN.04260413

18. Neri L, Kreuzberg U, Bellocchio F, et al. Detecting high-risk chronic kidney disease-mineral bone disorder phenotypes among patients on dialysis: a historical cohort study. Nephrol Dial Transplant. Apr 1 2019;34(4):682–691. doi:10.1093/ndt/gfy273

19. Stevens LA, Djurdjev O, Cardew S, Cameron EC, Levin A. Calcium, phosphate, and parathyroid hormone levels in combination and as a function of dialysis duration predict mortality: evidence for the complexity of the association between mineral metabolism and outcomes. J Am Soc Nephrol. Mar 2004;15(3):770–9. doi:10.1097/01.asn.0000113243.24155.2f

20. Kidney Disease: Improving Global Outcomes CKDMBD Working Group. KDIGO clinical practice guideline for the diagnosis, evaluation, prevention, and treatment of Chronic Kidney Disease-Mineral and Bone Disorder (CKD-MBD). Kidney Int Suppl. Aug 2009;(113):S1–130. doi:10.1038/ki.2009.188

21. Kidney Disease: Improving Global Outcomes CKDMBD Update Working Group. KDIGO 2017 Clinical Practice Guideline Update for the Diagnosis, Evaluation, Prevention, and Treatment of Chronic Kidney Disease-Mineral and Bone Disorder (CKD-MBD). Kidney Int Suppl. Jul 2017;7(1):1–59. doi:10.1016/j.kisu.2017.04.001

22. Danese MD, Halperin M, Lowe KA, Bradbury BD, Do TP, Block GA. Refining the definition of clinically important mineral and bone disorder in hemodialysis patients. Nephrol Dial Transplant. Aug 2015;30(8):1336–44. doi:10.1093/ndt/gfv034

23. Fishbane S, Shapiro WB, Corry DB, et al. Cinacalcet HCl and concurrent low-dose vitamin D improves treatment of secondary hyperparathyroidism in dialysis patients compared with vitamin D alone: the ACHIEVE study results. Clin J Am Soc Nephrol. Nov 2008;3(6):1718–25. doi:10.2215/CJN.01040308

24. Laurain E, Ayav C, Erpelding ML, et al. Targets for parathyroid hormone in secondary hyperparathyroidism: is a "one-size-fits-all" approach appropriate? A prospective incident cohort study. BMC Nephrol. Aug 13 2014;15(1):132. doi:10.1186/1471-2369-15-132

25. Messa P, Macario F, Yaqoob M, et al. The OPTIMA study: assessing a new cinacalcet (Sensipar/Mimpara) treatment algorithm for secondary hyperparathyroidism. Clin J Am Soc Nephrol. Jan 2008;3(1):36–45. doi:10.2215/CJN.03591006

26. Wetmore JB, Gurevich K, Sprague S, et al. A Randomized Trial of Cinacalcet versus Vitamin D Analogs as Monotherapy in Secondary Hyperparathyroidism (PARADIGM). Clin J Am Soc Nephrol. Jun 5 2015;10(6):1031–40. doi:10.2215/CJN.07050714

27. Ketteler M, Martin KJ, Wolf M, et al. Paricalcitol versus cinacalcet plus low-dose vitamin D therapy for the treatment of secondary hyperparathyroidism in patients receiving haemodialysis: results of the IMPACT SHPT study. Nephrol Dial Transplant. Aug 2012;27(8):3270–8. doi:10.1093/ndt/gfs018

28. Spiegel DM, McPhatter L, Allison A, Drumheller JC, Lockridge R. A computerized treatment algorithm trial to optimize mineral metabolism in ESRD. Clin J Am Soc Nephrol. Apr 2012;7(4):632–9. doi:10.2215/CJN.08170811

