## Supplemental Material for "Chronic Kidney Disease Mineral and Bone Disorder (CKD-MBD) Medication Titration Practices in Hemodialysis Patients in the US"

**Table of Contents:**

**Supplemental Methods**……………………………………………………..**2**

**Supplemental Tables**………………………………………………………..**8**

**Supplemental Figures**………………………………………………………**15**

**Supplemental Methods**

*Supplemental Information on Drug Dosing*

Presence and doses of medications were ascertained from the DCI Medication List. For each medication a small number of observations included doses that were deemed highly implausible and thus likely to be entry errors. These out-of-range values were recoded to missing doses. In addition, a few individuals used calcimimetics as part of clinical trials. These doses were reclassified as missing because identity of the calcimimetic (cinacalcet or etelcalcetide) and dose could not be determined.

| Medication | Out-of-range |
| --- | --- |
| Vitamin D Sterol (Paricalcitol Equivalent) | >100 mcg /dose |
| Cinacalcet | <15 mg or >270 mg/dose |
| Phosphorus Binders | |
| Calcium carbonate | >10,000mg/dose |
| Calcium acetate | >13,340mg/dose |
| Sevelamer hydrochloride | >16,000mg/dose |
| Sevelamer carbonate | >16,000mg/dose |
| Lanthanum carbonate | 15,000mg/dose |
| Sucroferric oxyhydroxide | >5,000mg/dose |
| Ferric citrate | >20g/dose |

Doses were converted to daily equivalents using the following weights. For these drugs, frequencies equal to or more frequent than every 4 hours or listed as prn or ‘comment’ were not converted to daily doses because they were deemed implausible, unlikely to be long term doses, or we could not determine the true frequency.

| Frequency Code | Description | Daily Dosage Coding (Doses/Day) |
| --- | --- | --- |
| QOW | Every other week | 0.07 |
| QWK, Q1W, QTUE, QTHU, QSUN, Q4D | Once/week | 0.14 |
| BIW | 2 times/week | 0.29 |
| TIW, QHD, Q3D | 3 times/week, | 0.43 |
|  | Every 3 days |  |
| QIW, NHD | 4 times/week | 0.57 |
| 5XW | 5 times/week | 0.71 |
| BID, Q12H | 2 times/day | 2 |
| TID, AC, PC | 3 times/day | 3 |
| QID | 4 times/day | 4 |
| QOD | Every other day | 0.5 |
| QD, QAM, QHS, QPM | Daily | 1 |
| 1X | One time only | Missing |
| CMT | See comments | Missing |
| PRN | As necessary | N/A |

For phosphorus binding medications, an ordinal variable was created to indicate doses based on high and low dosing. The following rules were used to indicate low and high doses:

| Generic name | Low range | High range |
| --- | --- | --- |
| Calcium carbonate | <3000 mg daily | ≥3000 mg daily |
| Calcium acetate | <3,335 mg daily | ≥3,335 mg daily |
| Sevelamer hydrochloride | <4,000 mg daily | ≥4,000 mg daily |
| Sevelamer carbonate | <4,000 mg daily | ≥4,000 mg daily |
| Lanthanum carbonate | <3000 mg daily  (500 mg, 750 mg or 1000 mg tablets) | ≥3000 mg daily |
| Sucroferric oxyhydroxide | <1500 mg daily (500 mg tablets) | ≥1500 mg daily (500 mg tablets) |
| Ferric citrate | <5 tablets daily (1g per tablet) | ≥5 tablets daily (1g per tablet) |

*Covariate Ascertainment*

Demographics included age at initiation of hemodialysis, sex, race (black, white, other) and ethnicity as ascertained on the Center for Medicare and Medicaid Services (CMS) Form 2728. Comorbidity included: the cause of kidney failure indicated as diabetes mellitus, hypertension, glomerulonephritis or other based on CMS Form 2728; diabetes mellitus status based on CMS Form 2728; and the overall burden of comorbidity adapted from a previously published weighted index by Liu *et al*^1^, however we did not incorporate Medicare claims in our index because we did not require Medicare as the primary payer in our cohort. Instead we only used comorbidities as ascertained on CMS Form 2728. Dialysis adequacy was the most recent single-pool Kt/V value, dialysate calcium was the most recently prescribed bath, and CKD-MBD laboratories included serum phosphorus, albumin-corrected serum calcium (i.e. serum calcium) and intact parathyroid hormone (PTH), each from the DCI EHR.

*Creating Laboratory Panel Data*

Monthly panel data were created for titration and initiation models according to the schematics below. Relevant CKD-MBD laboratories were selected for these models including lagged values of serum phosphorus and albumin-corrected serum calcium. We did not include lagged values of PTH because it is typically only drawn quarterly. Values of PTH >10,000 pg/ml, serum phosphorus >20 mg/dl and serum calcium >20 mg/dl were deemed unreliable and recoded as missing. Calcium was corrected for serum albumin according to the following equation: [0.8*(4.0 g/dl-serum albumin g/dl)] + serum calcium mg/dl.

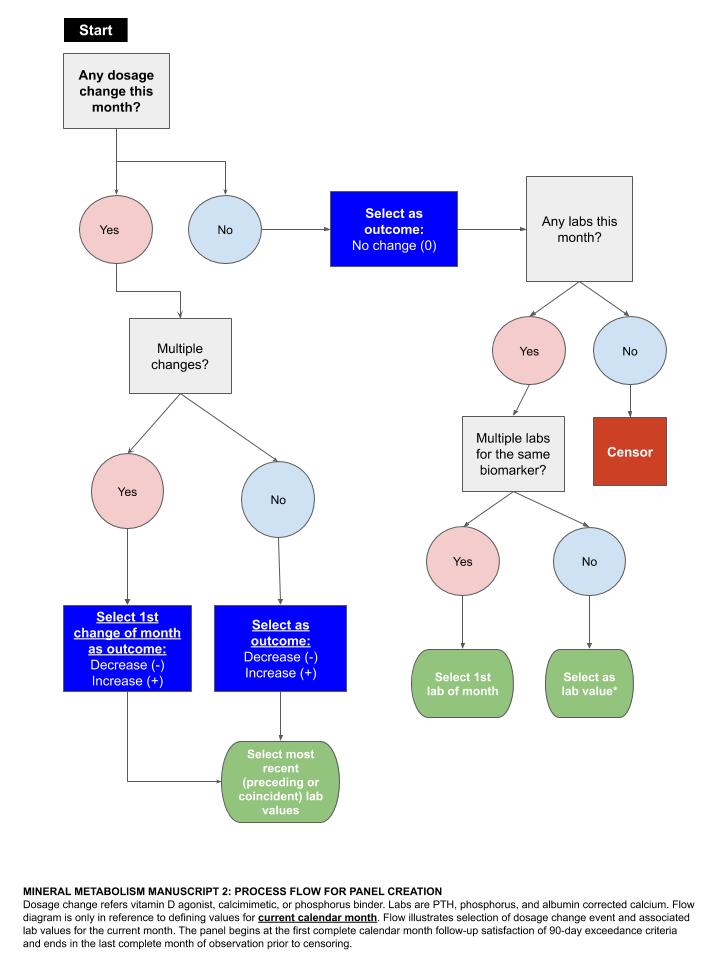

**Process for Panel Creation in the Current Month.** Dosage change refers to vitamin D sterols, calcimimetic, or phosphorus binder. Laboratories (labs) are parathyroid hormone (PTH), serum phosphorus, and albumin-corrected serum calcium. Flow diagram here references defining values for the current calendar month and depicts selection of dosage change event and associated lab values.

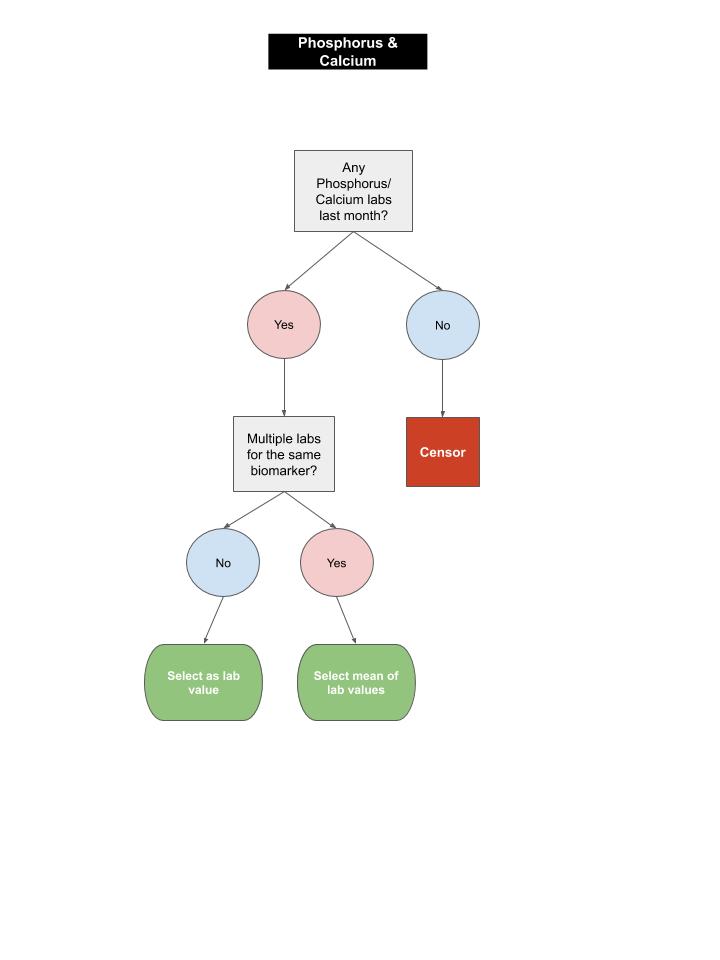

**Process for Panel Creation of Lagged Laboratories in the Prior Month.** Flow diagram here references defining values for the prior calendar month and depicts selection of individual or average values.

Lagged values of serum phosphorus and albumin-corrected calcium were incorporated to capture trends. To capture lags these were centered at the global mean, scaled by the standard deviation and then coded as the difference between the prior and current values to avoid collinearity with current measures already in the model. If more than one measure was available in a prior month, all measures were first averaged.

*Parameterization of Multivariable Models*

Patients could contribute person-months to models of vitamin D sterol titration and initiation, calcimimetic titration and initiation, and phosphorus binder titration and initiation based on having an eligible person-month in which they were (for titration models) or were not (for initiation models) using the drug of interest at the start of the month. Some individuals were not ultimately included in models because they did not have an eligible person-month with complete covariate data. This was usually due to lack of 3 months of lagged values for serum phosphorus and albumin-corrected serum calcium (**Supplemental Figure 1**).

We evaluated linear spline functions to model CKD-MBD laboratories. Candidate cutpoints were based on clinical practice guidelines, generally accepted laboratory reference ranges, and data distributions. The following cutpoints were evaluated either graphically and/or based on model fit: for PTH 150, 300, 400 and 600 pg/ml; for serum phosphorus 3.5, 5.5, and 7 mg/dl; and for serum calcium 8.0 and 10.2 mg/dl. We selected the final model with best fit according to Bayesian Information Criterion (BIC) for titration models. We evaluated selected patient characteristics including age categories (<30, 30-39, 40-49, 50-59, 60-64, 65-69, 70-74, 75-80, 80-84, ≥85 years), sex, diabetes, total comorbidity score (0-1, 2-3, 4-6, 7-9, ≥10) and functional limitation.

Final models for the **TITRATION** sample were parameterized as follows. A similar model was created for initiation models using mixed effects logistic regression.

(1) $ln\left( \frac{{Pr(Dose\_VitD}_{a,itf})}{{Pr(Dose\_VitD}_{0,itf})} \right)=\beta_{0}+\beta_{1}{cPTH}_{i,j}+\beta_{2}\left( {cPTH}_{i,j}-150 \right)_{+}+\beta_{3}\left( {cPTH}_{i,j}-250 \right)_{+}+\beta_{4}{cPhosphorus}_{i,j}+\beta_{5}\left( {cPhosphorus}_{i,j}-2 \right)_{+}+\beta_{6}{cCalcium}_{i,j}+\beta_{7}\left( {cCalcium}_{i,j}-2.2 \right)_{+}{+\beta}_{8}{({zPhos}_{i,j}-zPhos}_{i,j-1})+\beta_{9}{({zPhos}_{i,j}-zPhos}_{i,j-2})+\beta_{10}{({zPhos}_{i,j}-zPhos}_{i,j-3})+\beta_{11}{zCalcium}_{i,j}+\beta_{12}{({zCalcium}_{i,j}-zCalcium}_{i,j-1})+\beta_{13}{({zCalcium}_{i,j}-zCalcium}_{i,j-2})+\beta_{14}{({zCalcium}_{i,j}-zCalcium}_{i,j-3})+\vartheta_{i}+\delta_{f}+\varepsilon_{ij}$ ; where a = (-1,1)

(2) $ln\left( \frac{{Pr(Dose\_Calcimimetic}_{a,itf})}{{Pr(Dose\_Calcimimetic}_{0,itf})} \right)= \beta_{0}+\beta_{1}{cPTH}_{i,j}+\beta_{2}\left( {cPTH}_{i,j}-150 \right)_{+}+\beta_{3}\left( {cPTH}_{i,j}-250 \right)_{+}+\beta_{4}{cPhosphorus}_{i,j}+\beta_{5}\left( {cPhosphorus}_{i,j}-2 \right)_{+}+\beta_{6}{cCalcium}_{i,j}+\beta_{7}\left( {cCalcium}_{i,j}-2.2 \right)_{+}{+\beta}_{8}{({zPhos}_{i,j}-zPhos}_{i,j-1})+\beta_{9}{({zPhos}_{i,j}-zPhos}_{i,j-2})+\beta_{10}{({zPhos}_{i,j}-zPhos}_{i,j-3})+\beta_{11}{zCalcium}_{i,j}+\beta_{12}{({zCalcium}_{i,j}-zCalcium}_{i,j-1})+\beta_{13}{({zCalcium}_{i,j}-zCalcium}_{i,j-2})+\beta_{14}{({zCalcium}_{i,j}-zCalcium}_{i,j-3})+\vartheta_{i}+\delta_{f}+\varepsilon_{ij}$ ; where a = (-1, 1)

(3) $ln\left( \frac{{Pr(Dose\_Phos\_Binder}_{a,itf})}{{Pr(Dose\_Phos\_Binder}_{0,itf})} \right)= \beta_{0}+\beta_{1}{cPTH}_{i,j}+\beta_{2}\left( {cPTH}_{i,j}-150 \right)_{+}+\beta_{3}\left( {cPTH}_{i,j}-250 \right)_{+}+\beta_{4}{cPhosphorus}_{i,j}+\beta_{5}\left( {cPhosphorus}_{i,j}-2 \right)_{+}+\beta_{15}\left( {cPhosphorus}_{i,j}-3.5 \right)_{+}+\beta_{6}{cCalcium}_{i,j}+\beta_{7}\left( {cCalcium}_{i,j}-2.2 \right)_{+}{+\beta}_{8}{({zPhos}_{i,j}-zPhos}_{i,j-1})+\beta_{9}{({zPhos}_{i,j}-zPhos}_{i,j-2})+\beta_{10}{({zPhos}_{i,j}-zPhos}_{i,j-3})+\beta_{11}{zCalcium}_{i,j}+\beta_{12}{({zCalcium}_{i,j}-zCalcium}_{i,j-1})+\beta_{13}{({zCalcium}_{i,j}-zCalcium}_{i,j-2})+\beta_{14}{({zCalcium}_{i,j}-zCalcium}_{i,j-3})+\vartheta_{i}+\delta_{f}+\varepsilon_{ij}$; where a = (-1, 1)

Where ${Dose\_VitD}_{i,j}$, $Dose\_{Calcimimetic}_{i,j}$, and ${Dose\_PhosBinder}_{i,j}$ take the value of -1 when dosage is decreased or stopped, 0 if dosage doesn’t change and 1 if dosage is increased for vitamin D sterols (intravenous or oral), calcimimetics, and phosphorus binders (respectively) for person i in facility f at time point t. ${Calcium}_{i,j-k}$ and ${Phos}_{i,j-k}$ are laboratory values of calcium and phosphorus (respectively) at time j-k (where k=1,….,3 months); $\delta_{f}$ is a random intercept for facility f.

Values of PTH, phosphorus, and albumin corrected calcium are adjusted to be centered around the cutoff between “normal” and “low” values for each lab (150, 3.5, and 8.0 respectively). This is represented with the prefix of “c” (i.e. cPTH, cPhosphorus, cCalcium). The prefix of “z” indicates that these variables were z-scored.

Change points for the piecewise model were then coded with functions of the following form:

(Lab – Lab*)_+_ where “Lab” is the continuous value for the lab and Lab* is the chosen scalar change point value. The + indicates that the variable may only take a positive value and is set to 0 if the difference between the continuous lab value and the change point is ≤ 0. For example, if an individual had an observed PTH value of 200, then cPTH = 50, cPTH-150= -100, and cPTH-250=-200. Therefore, the value of the functions $\left( {cPTH}_{i,j}-150 \right)_{+}$ and $\left( {cPTH}_{i,j}-250 \right)_{+}$are both zero. Likewise, a PTH of 350 means cPTH is 200 and so the value of the first and second function are 50 and 0 respectively.

*Evaluation of Titration and Initiation Model Fit*

We qualitatively assessed model fit by comparing mean conditional predicted probabilities for the sample compared to conditional actual probabilities. To achieve this we first rounded laboratories into bands (nearest 75 pg/ml for PTH; 0.5 mg/dl for serum phosphorus and nearest 1.0 for albumin-corrected serum calcium). We calculated the mean predicted probabilities for patients with labs at selected points and graphed the probability. We then calculated the exact conditional probability at each point using the observed frequency and calculated 95% binomial exact confidence intervals. We plotted the predicted and observed with 95% confidence bands for visual comparison. For better stability of estimated exact probabilities, we required at least 20 observations within each range of lab values.

**Supplemental Tables**

**Supplemental Table 1. Characteristics of the Study Populations Included in Initiation Models.**

| Median, interquartile range or n(%) | Calcimimetic (N=18,153) | Phosphorus Binder (N=7,454) | Vitamin D Sterol (N=15,958) |
| --- | --- | --- | --- |
| **Age at incidence (years)** |  |  |  |
|  | 63 | 66 | 63 |
|  | 53, 73 | 56, 76 | 53, 73 |
| **Race** |  |  |  |
| White | 10,805 (59.5%) | 4,320 (58.0%) | 9,642 (60.4%) |
| Black/African American | 6,475 (35.7%) | 2,826 (37.9%) | 5,531 (34.7%) |
| Other | 873 (4.8%) | 308 (4.1%) | 785 (4.9%) |
| **Sex** |  |  |  |
| Male | 10,304 (56.8%) | 4,041 (54.2%) | 8,995 (56.4%) |
| Female | 7,849 (43.2%) | 3,413 (45.8%) | 6,963 (43.6%) |
| **Hispanic ethnicity** |  |  |  |
| No | 17,004 (93.7%) | 6,997 (93.9%) | 14,964 (93.8%) |
| Yes | 1,139 (6.3%) | 451 (6.1%) | 986 (6.2%) |
| **Primary cause of kidney failure** |  |  |  |
| Diabetes | 8,505 (50.6%) | 3,347 (48.8%) | 7,436 (50.4%) |
| Hypertension | 4,925 (29.3%) | 2,156 (31.4%) | 4,282 (29.0%) |
| Glomerulonephritis | 1,411 (8.4%) | 523 (7.6%) | 1,283 (8.7%) |
| Other cause | 1,954 (11.6%) | 833 (12.1%) | 1,753 (11.9%) |
| **Comorbidity index using medical evidence form (CMS Form 2728)** | | |  |
|  | 3 | 3 | 3 |
|  | 1, 5 | 1, 5 | 1, 5 |
| **Previously diagnosed with diabetes (CMS Form 2728)** | |  |  |
| No | 7,383 (40.7%) | 3,131 (42.1%) | 6,546 (41.1%) |
| Yes | 10,746 (59.3%) | 4,311 (57.9%) | 9,394 (58.9%) |
| **Inability to ambulate** |  |  |  |
| No | 16,176 (93.3%) | 6,567 (92.4%) | 14,191 (93.2%) |
| Yes | 1,157 (6.7%) | 542 (7.6%) | 1,035 (6.8%) |
| **Parathyroid hormone (90 days; pg/ml)** |  |  |  |
|  | 240 | 217 | 228 |
|  | 132, 422 | 120, 377 | 124, 413 |
| **Phosphorus (90 days; mg/dL)** |  |  |  |
|  | 5.2 | 4.8 | 5.2 |
|  | 4.3, 6.3 | 4.0, 5.7 | 4.3, 6.3 |
| **Albumin corrected calcium (90 days; mg/dL)** |  |  |  |
|  | 9.3 | 9.3 | 9.3 |
|  | 8.9, 9.7 | 8.9, 9.7 | 8.9, 9.7 |
| **Serum albumin (90 days; g/dL)** |  |  |  |
|  | 3.7 | 3.6 | 3.7 |
|  | 3.3, 3.9 | 3.3, 3.9 | 3.3, 3.9 |
| **Dialysis time (90 days; min/week)** |  |  |  |
|  | 630 | 630 | 630 |
|  | 630, 720 | 630, 720 | 630, 720 |
| **Kt/V (90 days)** |  |  |  |
|  | 1.5 | 1.5 | 1.5 |
|  | 1.4, 1.7 | 1.4, 1.7 | 1.4, 1.7 |
| **Dialysate calcium (90 days; mEq/L)** |  |  |  |
|  | 2.5 | 2.5 | 2.5 |
|  | 2.5, 2.5 | 2.5, 2.5 | 2.5, 2.5 |
| **Reason for censoring** |  |  |  |
| Transplant | 1,272 (7.0%) | 342 (4.6%) | 1,093 (6.8%) |
| Death | 7,205 (39.7%) | 3,410 (45.7%) | 6,391 (40.0%) |
| Transfer out of facility | 2,963 (16.3%) | 1,066 (14.3%) | 2,479 (15.5%) |
| End of observation period | 6,713 (37.0%) | 2,636 (35.4%) | 5,995 (37.6%) |

**Supplemental Table 2. Person-Months Reflecting Different Titration and Initiation Decisions**

|  | Calcimimetic (N=95,356) | Phosphorus Binders (N=426,891) | Vitamin D (N=263,093) |
| --- | --- | --- | --- |
| **Drug Titration Decision** |  |  |  |
| De-esclation | 4,499 (4.7%) | 12,327 (2.9%) | 45,653 (17.4%) |
| No Change | 87,170 (91.4%) | 398,903 (93.4%) | 184,194 (70.0%) |
| Escalation | 3,687 (3.9%) | 15,661 (3.7%) | 33,246 (12.6%) |

|  | Calcimimetic (N=420,785) | Phosphorus Binders (N=80,502) | Vitamin D (N=252,113) |
| --- | --- | --- | --- |
| **Drug Initiation** |  |  |  |
| No | 414,174 (98.4%) | 74,069 (92.0%) | 226,728 (89.9%) |
| Yes | 6,611 (1.6%) | 6,433 (8.0%) | 25,385 (10.1%) |

**Supplemental Table 3. Current and Lagged Laboratory Values by Status of Initiation for Vitamin D Sterols, Calcimimetics, and Phosphorus Binders**

| Mean ± SD | 1. Vitamin D  Started Drug this Month | | 2. Calcimimetic  Started Drug this Month | | 3. Phosphorus Binders  Started Drug this Month | |
| --- | --- | --- | --- | --- | --- | --- |
|  | No | Yes | No | Yes | No | Yes |
| PTH result | 328 ± 328 | 512 ± 332 | 322 ± 239 | 676 ± 393 | 308 ± 259 | 346 ± 293 |
| Phosphorus (mg/dl; current) | 5.5 ± 1.8 | 5.5 ± 1.4 | 5.4± 1.6 | 6.2 ± 1.8 | 4.7 ± 1.4 | 6.2 ± 1.6 |
| Change from -1 month | 0.0 ± 1.4 | -0.6 ± 1.5 | 0.0 ± 1.4 | 0.2 ± 1.4 | 0.1 ± 1.2 | 1.0 ± 1.6 |
| Change from -2 months | 0.0 ± 1.6 | -0.4 ± 1.6 | 0.0 ± 1.5 | 0.3 ± 1.6 | 0.1 ± 1.3 | 1.2 ± 1.8 |
| Change from -3 months | 0.0 ± 1.6 | -0.3 ± 1.7 | 0.0 ± 1.6 | 0.3 ± 1.7 | 0.1 ± 1.3 | 1.2 ± 1.8 |
| Albumin Corrected Calcium (mg/dl; current) | 9.2 ± 0.7 | 9.0 ± 0.7 | 9.2 ± 0.7 | 9.4 ± 0.7 | 9.2 ± 0.7 | 9.1 ± 0.8 |
| Change from -1 month | 0.0 ± 0.5 | -0.2 ± 0.6 | 0.0 ± 0.5 | 0.1 ± 0.6 | 0.0 ± 0.5 | -0.1 ± 0.6 |
| Change from -2 months | 0.0 ± 0.6 | -0.2 ± 0.7 | 0.0 ± 0.6 | 0.1 ± 0.7 | 0.0 ± 0.6 | -0.1 ± 0.7 |
| Change from -3 months | 0.0 ± 0.6 | -0.2 ± 0.7 | 0.0 ± 0.6 | 0.1 ± 0.7 | 0.0 ± 0.6 | -0.1 ± 0.7 |

**Supplemental Table 4. Selected Aikake’s and Bayesian Information Criteria Across Models During Model Building**

| **Model** | **AIC** | **BIC** |
| --- | --- | --- |
| **Vitamin D Sterols** |  |  |
| Laboratories and vintage | 335633 | 335785 |
| plus baseline patient characteristics | 335572 | 335875 |
| **Calcimimetic** |  |  |
| Laboratories and vintage | 63164 | 63298 |
| plus baseline patient characteristics | 63148 | 63416 |
| **Phosphorus Binders** |  |  |
| Laboratories and vintage | 212633 | 212803 |
| plus baseline patient characteristics | 212243 | 212572 |

| **Supplemental Table 5. Mixed Effects Multinomial Logistic Regression Model Estimates for Relative Odds of Escalation or De-escalation Relative to No Change for Each CKD-MBD Medication as a Function of CKD-MBD Laboratories** | | | | | | |
| --- | --- | --- | --- | --- | --- | --- |
|  | *Calcimimetic* | | *Phosphorus Binders* | | *Vitamin D Sterols* | |
| *Variable* | *Escalation*  *OR (95% CI)* | *De-escalation*  *OR (95% CI)* | *Escalation*  *OR (95% CI)* | *De-escalation*  *OR (95% CI)* | *Escalation*  *OR (95% CI)* | *De-escalation*  *OR (95% CI)* |
| **Original Regression Model Parameters** |  |  |  |  |  |  |
| Month | 0.993 ( 0.992, 0.995) | 0.997 ( 0.995, 0.998) | 0.990 ( 0.989, 0.991) | 0.997 ( 0.996, 0.998) | 0.992 ( 0.991, 0.993) | 0.999 ( 0.998, 0.999) |
| PTH (in 100s)-1.5 | 1.265 ( 1.035, 1.546) | 0.392 ( 0.370, 0.415) | 0.956 ( 0.930, 0.983) | 0.869 ( 0.844, 0.895) | 4.461 ( 4.177, 4.764) | 0.313 ( 0.307, 0.320) |
| PTH (Knot 300) | 6.222 ( 4.175, 9.272) | 3.192 ( 2.702, 3.770) | 0.929 ( 0.862, 1.000) | 1.032 ( 0.948, 1.123) | 0.793 ( 0.718, 0.876) | 4.633 ( 4.376, 4.904) |
| PTH (Knot 400) | 0.134 ( 0.106, 0.169) | 0.818 ( 0.725, 0.924) | 1.128 ( 1.067, 1.193) | 1.106 ( 1.035, 1.183) | 0.290 ( 0.277, 0.304) | 0.690 ( 0.662, 0.720) |
| Phosphorus-3.5 | 1.281 ( 1.199, 1.369) | 0.886 ( 0.849, 0.925) | 1.200 ( 1.156, 1.246) | 0.343 ( 0.334, 0.352) | 1.145 ( 1.122, 1.170) | 1.093 ( 1.073, 1.114) |
| Phosphorus (Knot 5.5) | 0.793 ( 0.733, 0.858) | 1.168 ( 1.102, 1.239) | 1.723 ( 1.616, 1.836) | 7.930 ( 7.429, 8.464) | 0.619 ( 0.600, 0.638) | 1.671 ( 1.631, 1.712) |
| Albumin Corrected Calcium – 8 | 1.288 ( 1.214, 1.367) | 0.723 ( 0.686, 0.762) | 1.057 ( 1.027, 1.087) | 1.127 ( 1.089, 1.167) | 0.852 ( 0.831, 0.873) | 1.638 ( 1.599, 1.677) |
| Phosphorus (Knot 7.0) | -- | -- | 0.437 ( 0.417, 0.459) | 0.384 ( 0.358, 0.411) | -- | -- |
| Albumin Corrected Calcium (Knot 10.2) | 0.841 ( 0.660, 1.071) | 2.614 ( 2.186, 3.125) | 2.427 ( 2.209, 2.667) | 1.929 ( 1.743, 2.135) | 0.184 ( 0.133, 0.256) | 2.760 ( 2.503, 3.044) |
| (Current-L1) Phosphorus | 0.975 ( 0.927, 1.025) | 0.975 ( 0.931, 1.022) | 0.967 ( 0.945, 0.990) | 0.997 ( 0.970, 1.025) | 1.050 ( 1.027, 1.074) | 1.072 ( 1.052, 1.094) |
| (Current-L1) Albumin Corrected Calcium | 0.996 ( 0.950, 1.045) | 0.938 ( 0.902, 0.977) | 0.960 ( 0.936, 0.985) | 0.978 ( 0.950, 1.006) | 0.861 ( 0.842, 0.881) | 1.197 ( 1.173, 1.221) |
| (Current-L2) Phosphorus | 1.064 ( 1.012, 1.118) | 0.993 ( 0.948, 1.039) | 1.099 ( 1.073, 1.125) | 0.906 ( 0.881, 0.931) | 0.982 ( 0.962, 1.003) | 0.992 ( 0.974, 1.011) |
| (Current-L2) Albumin Corrected Calcium | 1.034 ( 0.987, 1.083) | 0.963 ( 0.925, 1.002) | 0.949 ( 0.925, 0.973) | 0.999 ( 0.971, 1.028) | 0.899 ( 0.880, 0.918) | 1.095 ( 1.075, 1.116) |
| (Current-L3) Phosphorus | 1.090 ( 1.040, 1.142) | 0.932 ( 0.893, 0.973) | 1.214 ( 1.187, 1.242) | 0.960 ( 0.935, 0.985) | 1.007 ( 0.988, 1.027) | 0.956 ( 0.939, 0.972) |
| (Current-L3) Albumin Corrected Calcium | 0.993 ( 0.951, 1.036) | 0.980 ( 0.944, 1.018) | 0.968 ( 0.946, 0.991) | 1.012 ( 0.986, 1.040) | 0.881 ( 0.864, 0.899) | 1.108 ( 1.089, 1.128) |
| **Model Estimated Slopes at Various Lab Levels** |  |  |  |  |  |  |
| PTH < 300 (per 100 pg/ml) | 1.265 ( 1.035, 1.546) | 0.392 ( 0.370, 0.415) | 0.956 ( 0.930, 0.983) | 0.869 ( 0.844, 0.895) | 4.461 ( 4.177, 4.764) | 0.313 ( 0.307, 0.320) |
| PTH 300 to 400 (per 100 pg/ml) | 7.870 ( 6.249, 9.913) | 1.252 ( 1.112, 1.409) | 0.888 ( 0.842, 0.936) | 0.896 ( 0.842, 0.954) | 3.536 ( 3.377, 3.703) | 1.452 ( 1.395, 1.511) |
| PTH >400 (per 100 pg/ml) | 1.054 ( 1.048, 1.061) | 1.024 ( 1.016, 1.033) | 1.002 ( 0.995, 1.009) | 0.992 ( 0.982, 1.001) | 1.026 ( 1.021, 1.031) | 1.002 ( 0.996, 1.009) |
| Phosphorus ≤5.5 (per 1 mg/dl) | 1.281 ( 1.199, 1.369) | 0.886 ( 0.849, 0.925) | 1.200 ( 1.156, 1.246) | 0.343 ( 0.334, 0.352) | 1.145 ( 1.122, 1.170) | 1.093 ( 1.073, 1.114) |
| Phosphorus >5.5 (per 1 mg/dl) | 1.016 ( 0.986, 1.046) | 1.035 ( 1.003, 1.068) | 2.067 ( 1.994, 2.143) | 2.723 ( 2.595, 2.857) | 0.709 ( 0.696, 0.722) | 1.827 ( 1.802, 1.851) |
| Phosphorus >7.0 (per 1 mg/dl) | -- |  | 0.904 ( 0.884, 0.924) |  | -- |  |
| Calcium ≤10.2 (per 1 mg/dl) | 1.288 ( 1.214, 1.367) | 0.723 ( 0.686, 0.762) | 1.057 ( 1.027, 1.087) | 1.127 ( 1.089, 1.167) | 0.852 ( 0.831, 0.873) | 1.638 ( 1.599, 1.677) |
| Calcium >10.2 (per 1 mg/dl) | 1.083 ( 0.864, 1.357) | 1.890 ( 1.600, 2.233) | 2.565 ( 2.347, 2.802) | 2.174 ( 1.981, 2.386) | 0.157 ( 0.113, 0.217) | 4.520 ( 4.127, 4.951) |
| Groups (Patients) | 5,179 |  | 17,190 |  | 12,889 |  |
| Groups (Facilities) | 497 |  | 1,286 |  | 881 |  |
| Variance of Facility RE | 0.143 ( 0.108, 0.200) | 0.178 ( 0.136, 0.244) | 0.097 ( 0.078, 0.126) | 0.175 ( 0.138, 0.230) | 0.440 ( 0.364, 0.544) | 0.865 ( 0.709, 1.078) |
| Median Odds Ratio (MOR) | 1.433 (1.366, 1.529) | 1.493 (1.419, 1.599) | 1.345 (1.303, 1.402) | 1.488 (1.423, 1.577) | 1.878 (1.774, 2.015) | 2.419 (2.225, 2.682) |

***Note**: Due to linear spline terms and lagged laboratories, relationship of current laboratory values with escalation and de-escalation is most evident in ‘model estimated slopes’, which provides a linear combination of the spline estimates. Original estimates reflect changes in slope at the given knot. For instance, in the ‘original parameters’ the increasing odds of escalating calcimimetics with serum phosphorus that is higher than prior values 2 and 3 months ago is evident from the parameters for (Current-L2) Phosphorus and (Current-L3) Phosphorus, where L stands for ‘lagged’. However the association of increasing calcium values above 10.2 mg/dl on de-escalation of calcimimetics is best interpreted from the ‘model estimated slope’ for ‘calcium>10.2’. Intercept is not presented.

**Supplemental Table 6. Mixed Effects Logistic Regression Model Odds Ratio Estimates for Initiation for Each CKD-MBD Medication as a Function of Current CKD-MBD Laboratory Ranges**

|  | *Vitamin D Sterols* | *Calcimimetic* | *Phosphorus Binders* |
| --- | --- | --- | --- |
| **Model Estimated Slopes at Various Lab Levels** | *OR of Initiation (95% CI)* | | |
| PTH < 300 (per 100 pg/ml) | 4.13 ( 3.97, 4.30) | 1.36 ( 1.24, 1.49) | 0.97 ( 0.93, 1.02) |
| PTH 300 to 400 (per 100 pg/ml) | 2.89 ( 2.74, 3.04) | 9.93 ( 8.75, 11.26) | 1.05 ( 0.96, 1.15) |
| PTH >400 (per 100 pg/ml) | 0.98 ( 0.98, 0.99) | 1.16 ( 1.15, 1.17) | 1.02 ( 1.01, 1.03) |
| Phosphorus ≤5.5 (per 1 mg/dl) | 1.31 ( 1.27, 1.34) | 1.28 ( 1.23, 1.34) | 1.92 ( 1.81, 2.03) |
| Phosphorus >5.5 (per 1 mg/dl) | 0.56 ( 0.55, 0.58) | 1.21 ( 1.18, 1.24) | 2.48 ( 2.33, 2.64) |
| Phosphorus >7.0 (per 1 mg/dl) |  |  | 0.71 (0.68, 0.75) |
| Calcium ≤10.2 (per 1 mg/dl) | 0.93 ( 0.90, 0.95) | 2.10 ( 2.00, 2.21) | 0.96 ( 0.91, 1.01) |
| Calcium >10.2 (per 1 mg/dl) | 0.96 ( 0.82, 1.13) | 2.31 ( 2.00, 2.67) | 1.62 ( 1.36, 1.93) |

**Supplemental Table 7. Description of ‘Ceiling Doses’ at the Start of the Month by Titration Status for all Person-Months in Titration Models**

|  | *1. Vitamin D* | | | | | | *2. Calcimimetic* | | | | | | *3. Phosphorus Binders* | | | | | |
| --- | --- | --- | --- | --- | --- | --- | --- | --- | --- | --- | --- | --- | --- | --- | --- | --- | --- | --- |
|  | *De-escalation* | | *No change* | | *Escalation* | | *De-escalation* | | *No change* | | *Escalation* | | *De-escalation* | | *No change* | | *Escalation* | |
|  | *N* | *(%)* | *N* | *(%)* | *N* | *(%)* | *N* | *(%)* | *N* | *(%)* | *N* | *(%)* | *N* | *(%)* | *N* | *(%)* | *N* | *(%)* |
| *Drug at ceiling dose* | 98826 | 93.97 | 390726 | 96.48 | 77018 | 96.10 | 10202 | 94.94 | 177950 | 95.81 | 8842 | 98.09 | 20446 | 69.11 | 793374 | 86.82 | 39702 | 98.57 |
| *No* |  |  |  |  |  |  |  |  |  |  |  |  |  |  |  |  |  |  |
| *Yes* | 6342 | 6.03 | 14270 | 3.52 | 3126 | 3.90 | 544 | 5.06 | 7778 | 4.19 | 172 | 1.91 | 9140 | 30.89 | 120486 | 13.18 | 578 | 1.43 |

Note: Ceiling doses are defined as 120 mg daily for cinacalcet, 36 mcg weekly for paricalcitol equivalents, and 2 or more phosphorus binders at any dose at the start of the month. A few individuals are classified as at ‘ceiling doses’, yet escalating doses for phosphorus binders because these individuals escalated their dose on the first day of the month.

**Supplemental Figure 1. Flow of Patients into the Different Drug Titration and Initiation Models.** Data was modeled in panel format as person-months. Individuals may contribute person-months to both types of models based on their use of the drug at the beginning of the month. Sample size here represents unique individuals contributing person-months to each analysis.

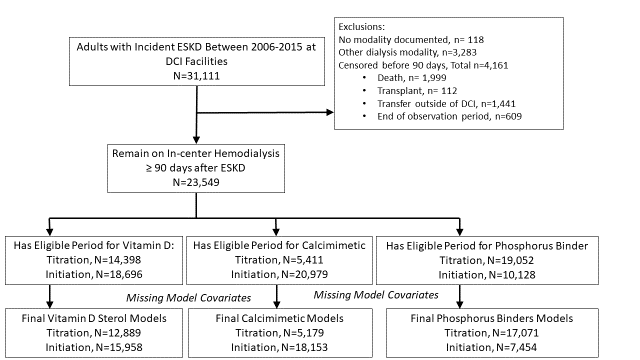

**References**

1.	Liu J, Huang Z, Gilbertson DT, Foley RN, Collins AJ. An improved comorbidity index for outcome analyses among dialysis patients. *Kidney Int.* 2010;77(2):141-151.
